# Health Risk Assessment for an Unregulated Neurotoxic Nicotine Analogue in Oral Pouch Products

**DOI:** 10.1101/2025.11.13.25340208

**Authors:** Sairam V. Jabba, Hanno C. Erythropel, Peter Silinski, Remi A. Mellinghoff, Paul T. Anastas, Julie B. Zimmerman, Sven E. Jordt

## Abstract

**Background:** Oral pouch products containing nicotine analogues such as 6-methyl nicotine (6MN) were recently introduced to evade federal and state regulations. Preclinical studies demonstrated that 6MN is more toxic than nicotine and binds to nicotinic receptors with higher potency. However, the nicotine analogue contents of oral pouch products and the associated health risks remain unknown.

**Methods:** Twenty-five flavor varieties from three oral pouch product (OPP) brands (Aroma-King, Hippotine-Happy Hippo, MG-Upperdeckys) marketed to contain 6MN were purchased in 2024-25. 6MN was quantified by either GC-FID (Yale) or GC-MS (Duke) to assess the estimated exposure dose (EED) from OPP use. Health risks associated with acute 6MN exposure from a single pouch use were assessed by calculating (i) hazard quotient (HQ) for heart-rate increase, and (ii) margin of exposure (MOE) for convulsions, with HQ more than 1 considered a potential health risk and the acceptable safety threshold value of MOE for proconvulsive effects considered 10. For comparison, both risk measures were determined for Zyn nicotine pouch products.

**Results:** 6MN contents in analyzed OPPs ranged from 2.96-14.5 mg and diverged significantly from labelled contents for some products. HQ’s for 6MN OPPs for heart-rate increase ranged from 74-381, compared to 95-162 for nicotine in Zyn. MOEs for convulsion risk ranged from 0.6-2.8, indicating elevated risk. MOEs for nicotine in Zyn products were 3.2-5.4.

**Conclusions:** 6MN contents in novel pouch products are higher than in popular nicotine pouch products. Consumers using these products are exposed to 6MN levels that may exceed safety thresholds, potentially leading to adverse health effects. Nicotine analogues should be urgently addressed by lawmakers and regulators, and FDA should be authorized to regulate products containing them.

## Introduction

The tobacco industry is constantly evolving, introducing new products with altered designs and constituents, to appeal to consumers and circumvent regulatory restrictions. Beginning in 2023, the industry introduced electronic-cigarettes and oral pouch products (OPPs), containing ‘unregulated’ nicotine analogues such as 6-methyl nicotine (6MN), in otherwise banned youth-appealing flavors.^1–3^ 6MN is more potent than nicotine at nicotinic receptors and in its neurotoxicity and lethality to rodents.^1–3^ This suggests that products containing 6MN have higher abuse potential and an elevated risk of adverse events. This study quantified 6MN in emerging OPPs and applied risk assessment methodologies to estimate the human health risks compared to products containing nicotine.

## Methods

### Product identification

“6-methyl nicotine” and variations thereof, and trade names for 6-methyl nicotine, including “Metatine”, “Nixotine”, “Nixodine”, “Imotine”, “Nonic6” and “Nonic” were chosen as web search terms to identify oral pouch products (OPPs) marketed within and outside of the US. Product labels and marketing information on vendor websites were reviewed for statements on product composition, including 6-methyl nicotine (6MN) and flavor chemicals. Product brands with tradenames Aroma King (Aroma King), Hippotine (Happy Hippo, LLC.) and MG (Upperdecky’s) were identified (25 flavor varieties in total) were purchased from web merchants in 2024-25 to confirm availability and for chemical analysis.

### Chemical Analysis

Contents of 6MN OPPs were extracted using methanol. As described in detail in Erythropel et al., 2024,^1^ 6MN was quantified by gas chromatography-flame ionization (GC-FID) at Yale University or gas chromatography-mass spectroscopy (GC-MS) at Duke University.^1^

### Risk Assessment

Health risks of acute 6MN exposure were assessed by (i)hazard quotient (**HQ**) for heart-rate increase, and (ii)margin of exposure (**MOE**) for convulsions, two measures widely applied by regulatory agencies to estimate human health risk from animal and human toxicological data. Calculated EEDs, HQs and MOEs were for a single pouch use (see Figure1A and eAppendix for details).

**Figure. 1.**
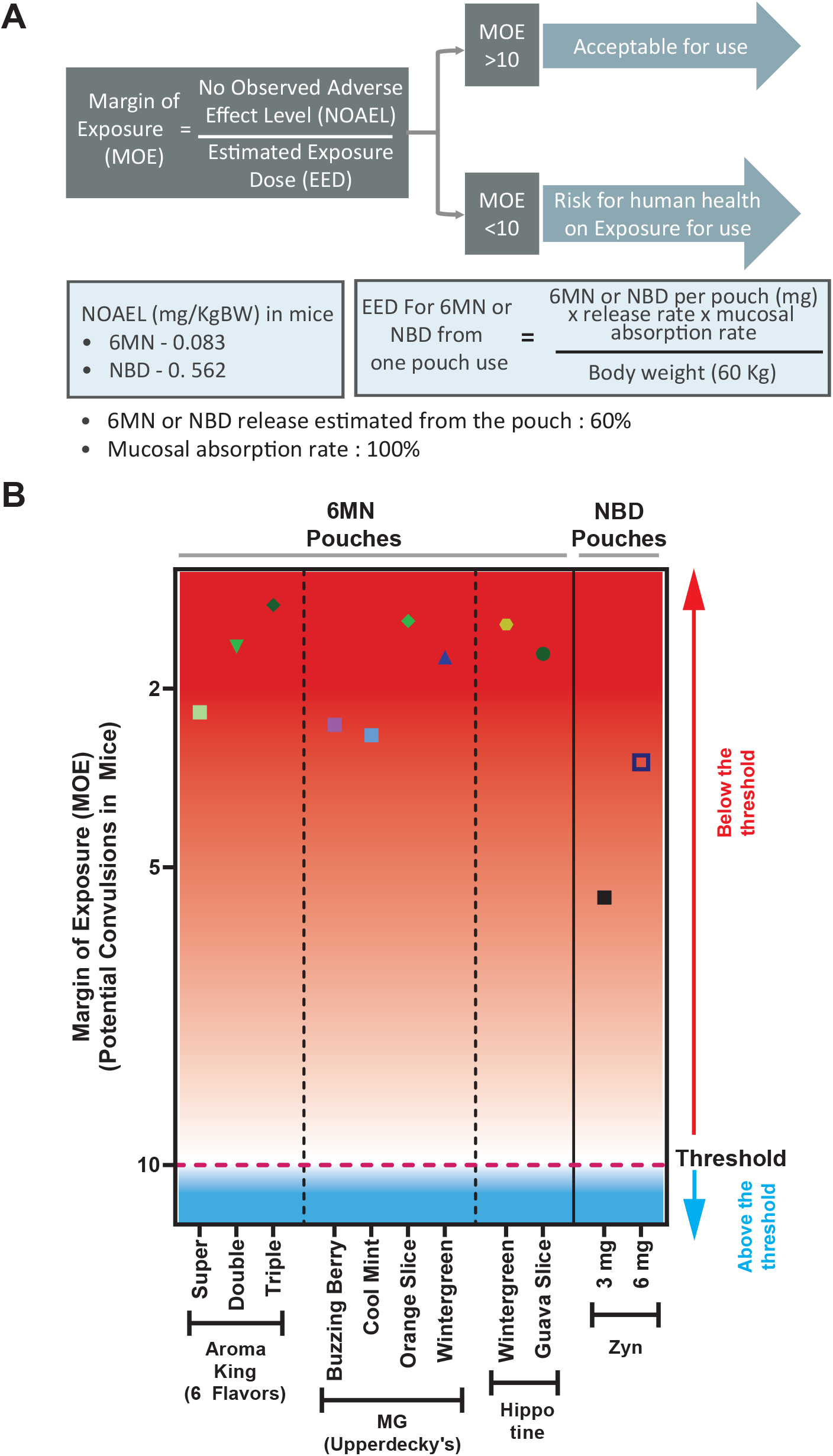
Margin of Exposure (MOE) Calculation Method and MOE Comparison for Various 6-Methyl Nicotine Oral Pouch Products. A) Calculation of MOE for neurotoxic chemical added to oral pouch products. An MOE of 10 was set as threshold for mitigation of potential proconvulsive activity of 6MN or nicotine. B) MOEs are plotted for 6MN exposures from use of one 6MN-containing OPP. Analyzed product brand tradenames included, Aroma King (Aroma King), Hippotine (Happy Hippo, LLC.) and MG (Upperdecky’s). MOEs determined for the popular nicotine-containing pouch product, Zyn, are displayed for comparison.

#### Hazard Quotient (HQ)

The risk assessment parameter named hazard quotient (HQ) was calculated to assess the acute cardiovascular health risk of increased heart rate associated with exposure to 6MN or nicotine from pouch use. HQ was determined as the ratio of exposure to 6MN (EED) to its acute reference dose for increases in heart rate, as follows:

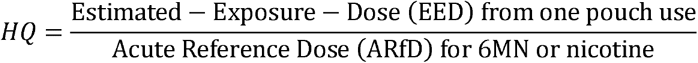

6MN’s ARfD for heart-rate increase, either in humans or preclinical animal models, is currently unavailable, but an ARfD of 0.8µg/KgBW has been established in humans for nicotine.^4–6^ This value is the external dose and upon correction for internal exposure (correction factor of 0.44 for oral bioavailability), an ARfD of 0.35µg/KgBW is obtained.^4,6,7^ As the physiochemical parameters of 6MN are similar to nicotine’s, as determined using computational toxicity toolbox (CompTox, EPA), a read-across approach with nicotine’s ARfD was utilized to calculate the ARfD of 6MN. Correcting for molecular weight (6MN: 176.23), the ARfD of 6MN was calculated to be 0.38µg/KgBW.

Estimated Exposure Dose (EED) is the estimated human exposure from use of a single 6MN or nicotine pouch and was calculated as follows:

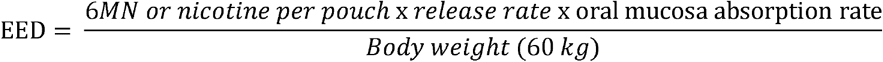

EED is the product of i) 6MN or nicotine bitartrate dihydrate (NBD) content per pouch, ii) their release rate from pouch to the oral mucosa, and iii) the absorption rate into the oral mucosa. With physicochemical properties of 6MN and nicotine being similar, release rate and absorption in oral mucosa from human pouch use for 6MN and nicotine were considered to be equivalent. A release rate of 60% and absorption rate through the oral mucosa of 100% was considered for these calculations.^4,7,8^ With EED’s represented on a ‘per Kilogram-Bodyweight (KgBW)’ basis, a body weight of 60 kg was assumed for these calculations. A HQ > 1 indicates a potential health risk (increased heart rate).

#### Margin of exposure (MOE)

To assess the health risk of convulsions associated with 6MN exposure from use of 6MN containing OPPs, we calculated MOE, a risk assessment parameter for toxic compounds widley used by regulatory agencies. For comparison with health risks from exposure to nicotine from similar products, MOE was calculated for use of a Zyn oral nicotine pouch (ONP; nicotine-reference products), in which nicotine bitartrate dihydrate (NBD) is the nicotine salt. For nicotine-reference ONPs, the label nicotine strength is based on the maximum amount of free-base nicotine released. The equivalent NBD amount added to a pouch was determined by correcting for NBD molecular weight (498.5) and utilizing a scaling factor of ∼3.07 (498.5 ÷ 162.2 = 3.07) to the amount of free-base nicotine determined in nicotine-reference pouches. This nicotine-reference product (Zyn) was selected for comparison because it is the most popular ONP currently in the market.

MOE was calculated using the following formula, and as described previously.^9,10^

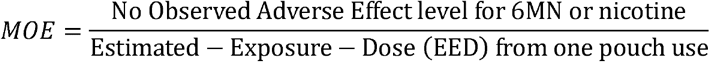

Here, NOAEL is the No-Observed-Adverse-Effect-Level, the highest administered dose to mice at which the adverse outcome of convulsions wasn’t observed. NOAELs of 6MN and NBD for convulsion were 82.5 and 562 µg/KgBW, respectively, derived from published studies determining the dose-response relationship to produce convulsions and lethality in mice.^11^ EED calculation was performed as described earlier.

The acceptable safety threshold value of MOE for proconvulsive chemicals was considered to be 10, with an uncertainty factor of 10 accounting for intra-species (within-human) variability. With rodent animal models (mice and rats) reported to have comparable convulsion risk as non-human primates and with similar sensitivity to humans for neurotoxic chemicals, an additional modifying factor for inter-species differences (rodent to human) was not applied.^12,13^ Therefore, an MOE of >10 is considered low risk, while MOEs <10 are potentially proconvulsive and require prioritization by regulatory agencies for risk mitigation.

## Results

Pouch 6MN contents ranged from 2.96-14.48mg. For products 2-3 measured 6MN contents were ∼61-64% lower than listed on the product label while for Product 1, no 6MN quantities were indicated on the label. Calculated HQs for heart-rate increase ranged from 74-381 for 6MN OPPs, compared to 95-162 for nicotine-containing products. Determined MOEs for convulsion risk ranged from 0.6-2.8 for 6MN OPPs, below the safety thresholds of 10. MOEs for nicotine-containing product were 3.2-5.5 (see Table and Figure1B).

## Discussion

This study demonstrates that US-marketed pouches contain 6MN at amounts (2.96-14.48mg/pouch) that exceed the typical nicotine content (2-8mg/pouch) in popular ONPs. Results are also comparable to EU- and UK-marketed OPPs found to contain 3.9-19.4 mg/pouch.^14^ Risk assessment based on 6MN contents and prior toxicological studies indicate that people who use just a single 6MN pouch are exposed to 6MN levels that exceed risk thresholds for adverse health effects, such as increased heart rate and convulsions.

Previous rodent toxicity studies demonstrated that 6MNs ED_50_ (110µg/KgBW) for convulsions and LD_50_ (190µg/KgBW) are ∼2-3X lower than those of nicotine (ED_50_=348µg/KgBW; LD_50_=422µg/KgBW),^11^ demonstrating that 6MN is significantly more toxic than nicotine. 6MN’s higher lipophilicity and divergent metabolism^15^ from nicotine may also contribute to greater bioaccumulation and toxicity.

Due to their similar physicochemical properties and pharmacological targets, the ARfD of 6MN was derived from that of nicotine, despite 6MN’s greater potency for adverse outcomes. Consequently, our analysis adopted a conservative approach that plausibly underestimated 6MNs health risks. Nevertheless, for all the products analyzed, both assessed risk measures indicate that exposures were above levels of concern.

Study limitations include testing only a limited number of brands.

Based on the acute health risks associated with 6MN intake from OPPs, the FDA and state regulators should consider restricting these products and assess their safety along with other 6MN-containing product categories, including e-cigarettes.

## Data Availability

All data produced in the present work are contained in the manuscript

## Disclaimer

The content is solely the responsibility of the authors and does not necessarily represent the views of the funding institute.

## Acknowledgements

The authors would like to thank Victor Garcia-Gallet for his assistance in OPP extraction.

## Author Contributions

Drs Jabba and Erythropel contributed equally. Drs Jabba and Jordt had full access to all of the data in the study and take responsibility for the integrity of the data and the accuracy of the data analysis.

**Concept and design:** Jabba, Jordt

**Acquisition, analysis, or interpretation of data:** Jabba, Erythropel, Silinski, Mellinghoff, Jordt

**Drafting of the manuscript:** Jabba, Jordt, Erythropel

**Critical revision of the manuscript for important intellectual content:** Jabba, Jordt, Erythropel, Anastas, Zimmerman.

**Supervision:** Jordt, Zimmerman.

**Obtained funding:** Zimmerman, Jordt.

**Administrative, technical, or material support:** Jabba, Jordt, Erythropel, Anastas, Zimmerman.

---

**Table.** Chemical Analysis and Predicted Risk Measures (HQ and MOE) for US-Marketed 6-Methyl Nicotine Containing Oral Pouch Products Tobacco.

| Brand Name | Flavor Name | Label Strength | 6MN or nicotine content, mean (SD) in mg/pouch, Difference from label strength [%] |  | Risk Assessment Parameter |  |
| --- | --- | --- | --- | --- | --- | --- |
|  |  |  | 6MN | Nicotine | HQ | MOE |
| Aroma King (Product 1) | Exotic Ice | Extra Strong | 3.28 (0.11) |  | 86 | 2.5 |
|  |  | Super Strong | 5.71 (0.15) |  | 150 | 1.4 |
|  |  | Mega Strong | 11.53 (0.17) |  | 303 | 0.7 |
|  | Cola Ice | Extra Strong | 3.76 (0.01) |  | 99 | 2.2 |
|  |  | Super Strong | 6.41 (0.04) |  | 169 | 1.3 |
|  |  | Mega Strong | 12.09 (0.26) |  | 318 | 0.7 |
|  | Rainbow Drops | Extra Strong | 3.26 (0.09) |  | 86 | 2.5 |
|  |  | Super Strong | 7.18 (0.07) |  | 189 | 1.1 |
|  |  | Mega Strong | 13.29 (0.14) |  | 350 | 0.6 |
|  | Freeze Ice<sup>*,#</sup> | Extra Strong | 3.49 (0.57) |  | 92 | 2.4 |
|  |  | Super Strong | 5.91 (0.74) |  | 156 | 1.4 |
|  |  | Mega Strong | 12.25 (0.81) |  | 322 | 0.7 |
|  | Peach Ice<sup>*,#</sup> | Extra Strong | 3.33 (0.22) |  | 88 | 2.5 |
|  |  | Super Strong | 6.05 (0.31) |  | 159 | 1.4 |
|  |  | Mega Strong | 13.01 (0.93) |  | 342 | 0.6 |
|  | Ruby Berry Ice<sup>*</sup> | Extra Strong | 3.46 (0.32) |  | 91 | 2.4 |
|  |  | Super Strong | 7.84 (0.77) |  | 206 | 1.1 |
|  |  | Mega Strong | 13.88 (0.85) |  | 365 | 0.6 |
|  | Double Mint | Mega Strong | 14.48 (0.85) |  | 381 | 0.6 |
| MG (Product 2) | Buzzing Berry<sup>*,#</sup> | 8 mg | 3.16 (0.30) [-61%] |  | 83 | 2.6 |
|  | Cool Mint<sup>*,#</sup> | 8 mg | 2.96 (0.25) [-63%] |  | 78 | 2.8 |
|  | Orange Creamsicle<sup>*,#</sup> | 25 mg | 9.53 (0.96) [-62%] |  | 251 | 0.9 |
|  | Wintergreen<sup>*,#</sup> | 15 mg | 5.59 (0.27) [-63%] |  | 147 | 1.5 |
| Hippotine (Product 3) | Guava Slice<sup>*,#</sup> | 15 mg | 5.81 (0.61) [-61%] |  | 153 | 1.4 |
|  | Wintergreen<sup>*,#</sup> | 25 mg | 8.90 (1.14) [-64%] |  | 234 | 0.9 |
| Zyn<sup>&</sup> (nicotine-reference products) | All Flavors<sup>§</sup> | 3 mg |  | 3.32 (0.11) [11%] | 95 | 5.5 |
|  |  | 6 mg |  | 5.66 (0.34) [-6%] | 162 | 3.2 |

\*N=5-7 for 6MN from 3 separate pouch extractions, remaining N=3 from one extraction

